# CNNeoPP: A Deep Learning Pipeline for Personalized Neoantigen Prediction and Liquid Biopsy Applications

**DOI:** 10.1101/2025.03.22.25324446

**Authors:** Yu Cai, Rui Chen, Mingming Song, Lei Wang, Zirong Huo, Dongyan Yang, Sitong Zhang, Shenghan Gao, Seungyong Hwang, Ling Bai, Yonggang Lv, Yali Cui, Xi Zhang

## Abstract

Neoantigens have emerged as promising targets for personalized cancer immunotherapy. However, accurate identification of immunogenic neoantigens remains a challenge due to limitations in existing predictive models. Here, we present CNNeo, a novel deep learning-based neoantigen prediction model, and CNNeoPP, an integrated computational pipeline for neoantigen discovery. CNNeo employs natural language processing-based sequence encoding and multi-modal feature integration, demonstrating superior predictive performance compared to existing tools. CNNeoPP was rigorously validated using independent datasets, including the TESLA dataset, and experimental validation via ELISpot T-cell assays. Additionally, we conducted a proof-of-concept study utilizing plasma cell-free DNA to explore the feasibility of non-invasive neoantigen prediction. We found that increased sequencing depth enhances neoantigen detectability, further amplified by the prioritization strategy of CNNeoPP. CNNeoDB, a publicly accessible database was developed compiling neoantigen data from multiple sources. This study establishes robust tools for neoantigen prediction, with implications for optimizing cancer immunotherapy and liquid biopsy-based tumor monitoring.

## Introduction

Neoantigens are tumor-specific antigens that arise due to somatic mutations in cancer cells or from viral proteins. These novel peptide sequences are presented by major histocompatibility complex (MHC) molecules and can be recognized by T lymphocytes^1^. Due to their high immunogenicity, exclusive expression in tumor cells, and poor tolerance by the immune system, neoantigens can elicit patient-specific immune responses, making them an attractive target for cancer immunotherapy^2,3^. Neoantigen-based therapies, including personalized vaccines and adoptive T cell therapies, have gained momentum in recent years, with rapid advancements accelerating their clinical applications^4,5^. Moreover, neoantigens are increasingly being utilized to assess responses to immunotherapy^6^, and their detection through liquid biopsy approaches, such as tumor cell-free DNA (cfDNA) analysis, offers a promising non-invasive strategy for monitoring tumor evolution and guiding treatment decisions^7,8^.

Somatic single nucleotide variants (SNVs), which account for approximately 80% of neoantigens^9^, can be efficiently detected using next-generation sequencing (NGS) technologies like whole-exome sequencing (WES) and RNA sequencing (RNA-seq). Despite reductions in NGS costs and advancements in sequencing technologies, neoantigen identification through NGS still faces challenges^1,10^. Many computational models predominantly rely on peptide-MHC binding affinity predictions^11^, often neglecting immunogenicity factors such as antigen processing, peptide stability, T cell receptor recognition, and immune regulation^12^. Traditional machine learning models and simplistic scoring methods fail to capture the complex sequence patterns underpinning immune recognition, limiting their predictive accuracy. To address these shortcomings, we have proposed deep learning-based approaches incorporating advanced sequence encoding techniques to enhance neoantigen prediction accuracy^10^. Additionally, validation of neoantigen prediction models has largely relied on *in silico* analyses or limited experimental data, lacking rigorous independent validation, highlighting the critical need for robust experimental validation. Furthermore, existing prediction models are predominantly optimized for bulk tumor tissue data, leaving their applicability to liquid biopsy approaches uncertain, despite the potential of cfDNA-based neoantigen identification to improve treatment strategies^13^.

Convolutional Neural Networks (CNNs) are a class of deep learning models designed to capture hierarchical features and spatial patterns within sequential or image-like data. Natural Language Processing (NLP), a subfield of artificial intelligence, processes textual or sequence-based data using tokenization and embedding techniques. To overcome limitations described above, we developed a deep learning-based neoantigen prediction model, CNNeo (“CNN and NLP-based Neoantigen Prediction Model”), optimized for performance by integrating neoantigen peptide sequences and human leukocyte antigen (HLA) allele information using NLP-based encoders. Additionally, we assembled and validated CNNeoPP (“CNNeo Pipeline”), a comprehensive pipeline integrating NGS data processing with neoantigen classification. These advancements facilitate neoantigen discovery from plasma cfDNA, supporting the development of personalized cancer immunotherapies.

## Results

### Study Design and Overall Strategy

This study (Fig. 1) details the development and validation of CNNeo, a novel deep learning-based neoantigen prediction model, and its integration into CNNeoPP, a comprehensive computational pipeline for neoantigen discovery. CNNeoPP incorporates sequencing data processing, mutation calling, feature extraction, and immunogenicity prediction using NGS data from tumor tissue and PBMCs. Independent validation was performed using the TESLA dataset, T-cell assays, and a cfDNA-based proof-of-concept study. Additionally, we developed CNNeoDB, a curated neoantigen database to facilitate data sharing and support future research.

**Figure 1.**
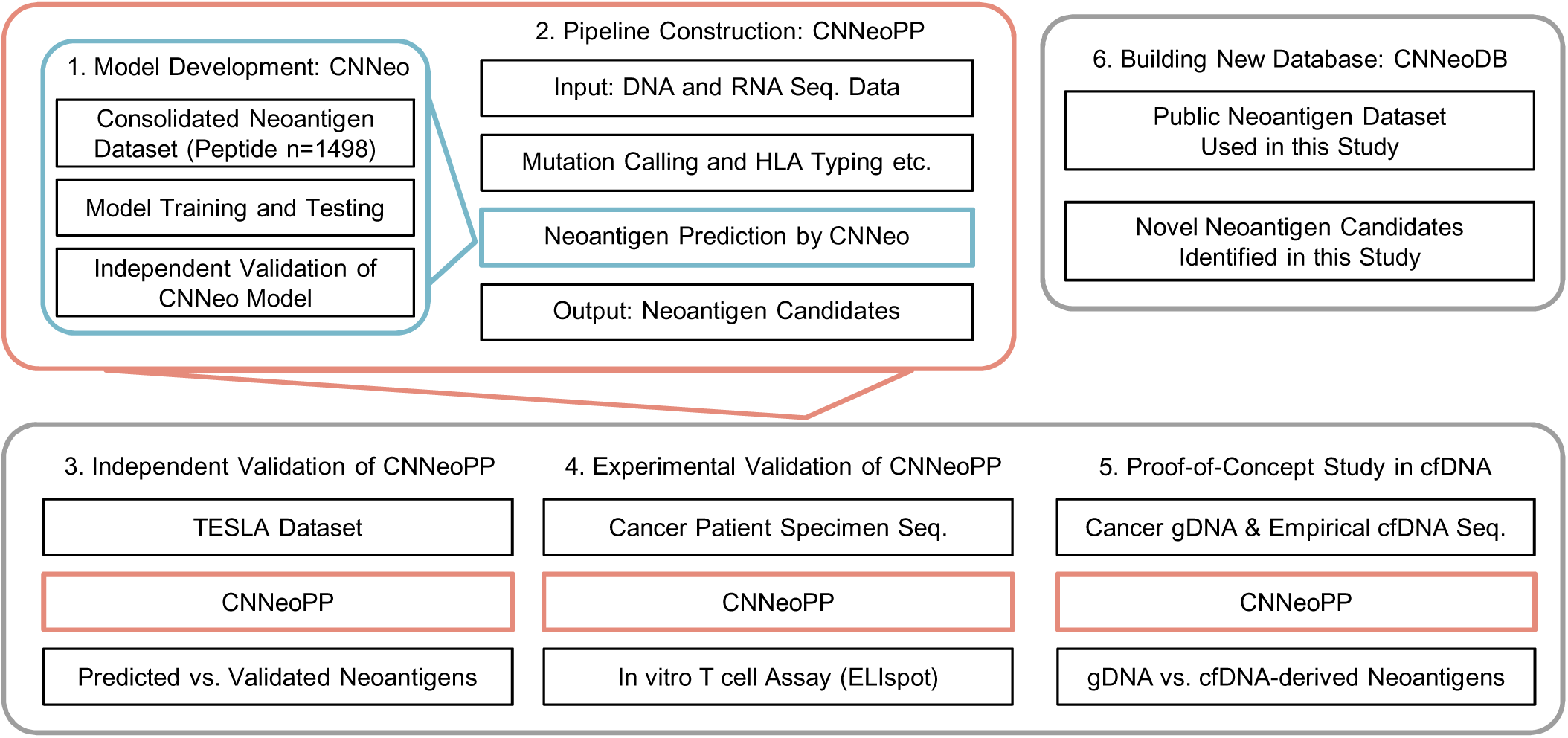
Overall Study Design. This study follows six key steps: (1) development of CNNeo, a neoantigen prediction model trained on immunogenic and non-immunogenic peptide datasets; (2) implementation of CNNeoPP, a computational pipeline integrating CNNeo for neoantigen prediction from sequencing data; (3) independent validation using the TESLA dataset; (4) experimental validation through ELISpot assays; (5) a proof-of-concept study comparing neoantigen identification from gDNA and cfDNA using CNNeoPP; and (6) development and release of CNNeoDB, a comprehensive neoantigen database integrating both public and newly identified candidates from this study.

### Data Collection and Feature Evaluation

The training dataset was curated from literature and public databases, including Neodb and NEPdb, yielding a dataset of 1,498 validated peptides (Fig. S1A, Table S1). An independent validation dataset consisting of 153 neoantigens was compiled (Fig. S1B). The TESLA dataset, which included paired tumor-normal samples and experimentally validated peptides, was used for further validation (Fig. S1C).

In the training dataset, immunogenic peptides exhibited distinct biological characteristics compared to non-immunogenic peptides, with a notable enrichment of shorter peptide lengths (9-mers being the most common) (Fig. 2A) and differences in amino acid composition (Figs. 2B & 2C). Key residues such as glutamic acid (E) and Aspartic acid (D) are more enriched in immunogenic peptides at positions P2, P3 and P4, and lysine (K), arginine (R), are predominantly seen at position P1. Analysis of HLA-I allele distribution revealed that immunogenic peptides were associated with a more diverse range of HLA alleles (Fig. S2). A statistical comparison of 11 immunogenicity-related features (Tables S2 & S3) showed significant differences in 7 features (Fig. S3), and most features remained independent (Fig. S4). Random forest analysis identified TAP transport efficiency, NetCTLpan score, and peptide-HLA binding affinity as the top 3 predictive features of immunogenicity (Fig. 2D).

**Figure 2.**
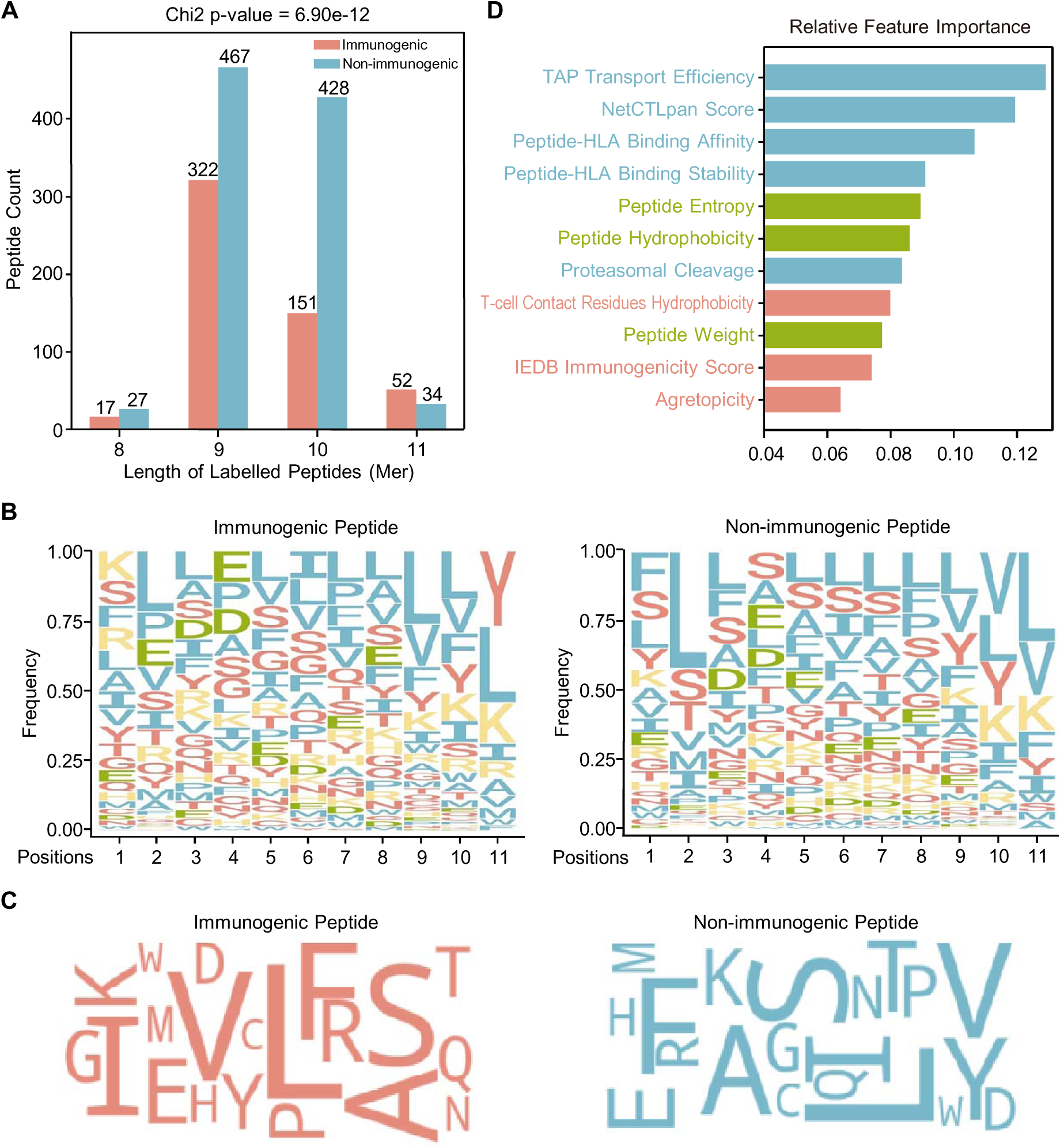
**Characterization of Peptides in the Training Dataset and Feature Importance**. (A) Peptide length distribution comparing immunogenic and non-immunogenic peptides. The statistical significance of differences in peptide length distributions was evaluated using a chi-square test, with the p-value displayed in the figure title. (B) Sequence logos representing amino acid preferences in immunogenic (left) and non-immunogenic (right) peptides. Amino acids are color-coded based on their properties: polar (red), nonpolar/hydrophobic (blue), positively charged (yellow), and negatively charged (green). The y-axis represents the normalized relative frequency of each amino acid, ensuring the sum equals 1. (C) Word clouds visualizing amino acid composition of immunogenic (left) and non-immunogenic (right) peptides. The relative size of each amino acid reflects its frequency within each group, illustrating compositional differences between immunogenic and non-immunogenic peptides. (D) Feature importance analysis using a random forest model. The relative contribution of 11 immunogenicity features is ranked to assess their impact on immunogenicity classification.

### Model Training, Testing and Independent Validation

A total of 24 individual models were trained utilizing a variety of supervised learning algorithms incorporating peptide sequences, HLA allele information, and immunogenicity features (Table S4, Fig. 3A), with three models outperforming the others (Fig. 3B). The FCNN_TF and CNN_BioBERT models demonstrated the highest performance, achieving an AUC of 0.81 by leveraging TF-IDF or BioBERT encoding to extract semantic relationships in peptide sequences. The FCNN_BioBERT model exhibited robustness by integrating deep learning with combined immunogenicity features, peptide sequences, and HLA features, improving predictive performance compared to models using only immunogenicity features. These three top-performing models were then integrated into the CNNeo framework and tested using an independent dataset of 69 immunogenic and 84 non-immunogenic mutant peptides (Fig. 3C). The CNNeo model, along with its individual components (FCNN_TF, CNN_BioBERT, and FCNN_BioBERT), was benchmarked against four existing tools (DeepImmuno-CNN, seq2neo-CNN, Immuno-GNN, and INeo-Epp). Despite prediction variability among models, performance evaluation at different ranking thresholds (top 10, top 20, and top 50) revealed that the CNNeo framework outperformed all existing tools (Fig. 3C, Table S5), further demonstrating the benefits of integrating combined features and algorithms.

**Figure 3.**
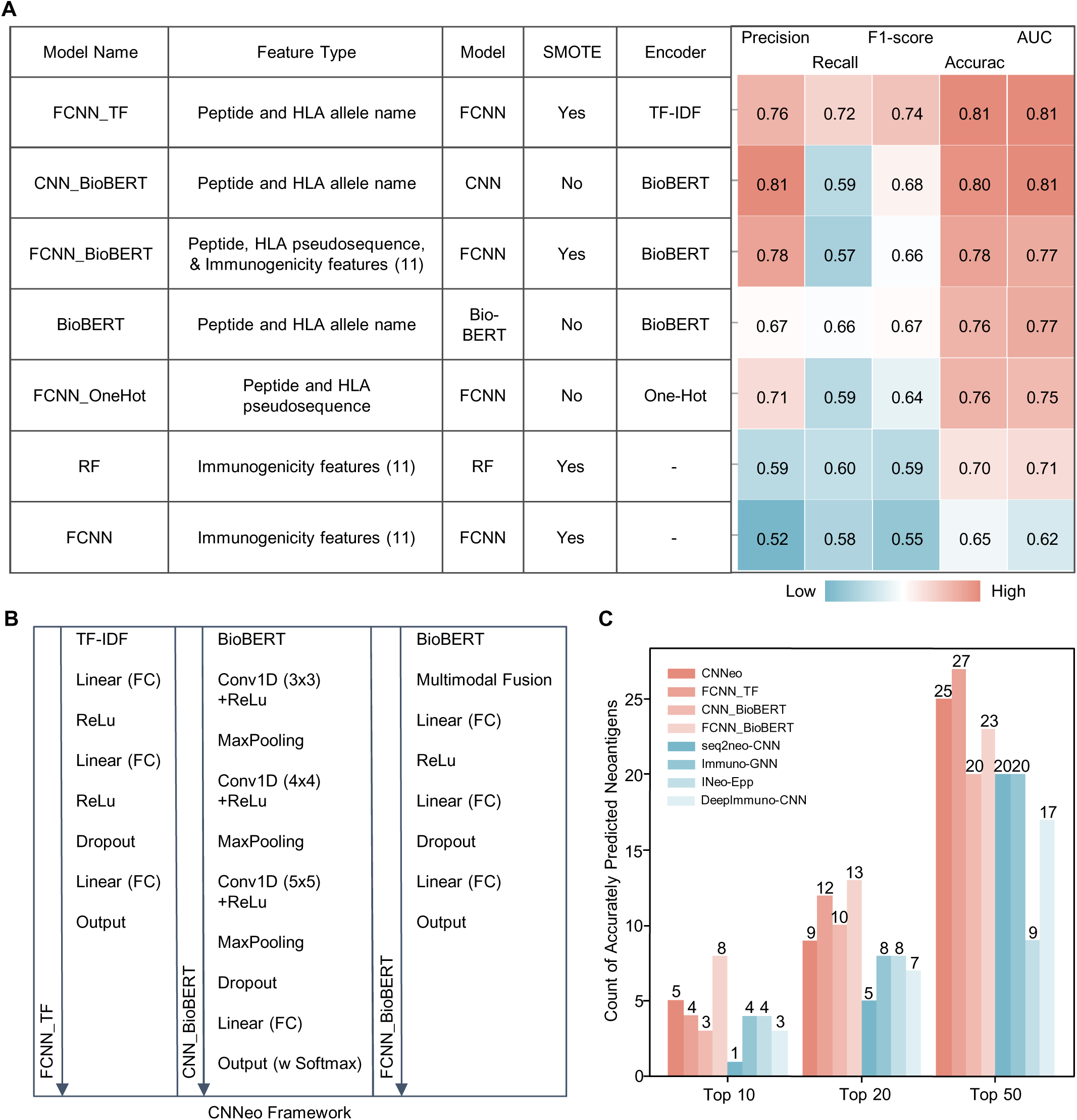
Architecture and Performance Evaluation of CNNeo. (A) Heatmap summarizing key model components (e.g., feature type, algorithm, encoding method) alongside performance metrics of representative models used in model development. Color transitions from teal to pink indicate increasing performance values across evaluation metrics. (B) Schematic representation of three top-performing neural network architectures integrated into the CNNeo framework, illustrating the model workflow from encoding method (top) to final predictions (bottom). (C) Comparative analysis of predicted immunogenic neoantigen counts across different models in TopN rankings using an independent dataset. CNNeo (integrating FCNN_TF, CNN_BioBERT, and FCNN_BioBERT) is compared against its individual components and four existing tools (DeepImmuno-CNN, seq2neo-CNN, Immuno-GNN, INeo-Epp), evaluating their relative predictive performance and the advantages of ensemble integration.

### Full Pipeline-Level Validation Using TESLA Dataset

CNNeo was then integrated into CNNeoPP, a comprehensive pipeline designed to streamline neoantigen discovery by performing DNA-seq on tumor tissue and PBMC samples, as well as RNA-seq on tumor tissue, followed by analyzing sequencing data, somatic SNVs, RNA expression levels, and HLA typing (Fig. 4A). The performance of CNNeoPP was further evaluated using the TESLA consortium dataset (Table S6) and compared with pipelines that integrate existing tools, including seq2neo-CNN, Immuno-GNN and INeo-Epp. Among 532 tested SNV-derived peptides from 5 patients in the TESLA dataset, 34 (6.39%) were experimentally confirmed as immunogenic, indicating that only a small fraction of somatic mutations generates neoantigens capable of eliciting a T-cell response (Fig. 4B). CNNeoPP successfully captured 3, 5, and 8 unique immunogenic peptides within the top 10, 20, and 50 candidates, respectively, demonstrating a clear and reproducible advantage over existing approaches (Fig. 4C, Table S7). This consistent performance underscores the robustness of the multi-model ensemble pipeline and its superior accuracy in identifying neoantigens in real-world and independent applications.

**Figure 4.**
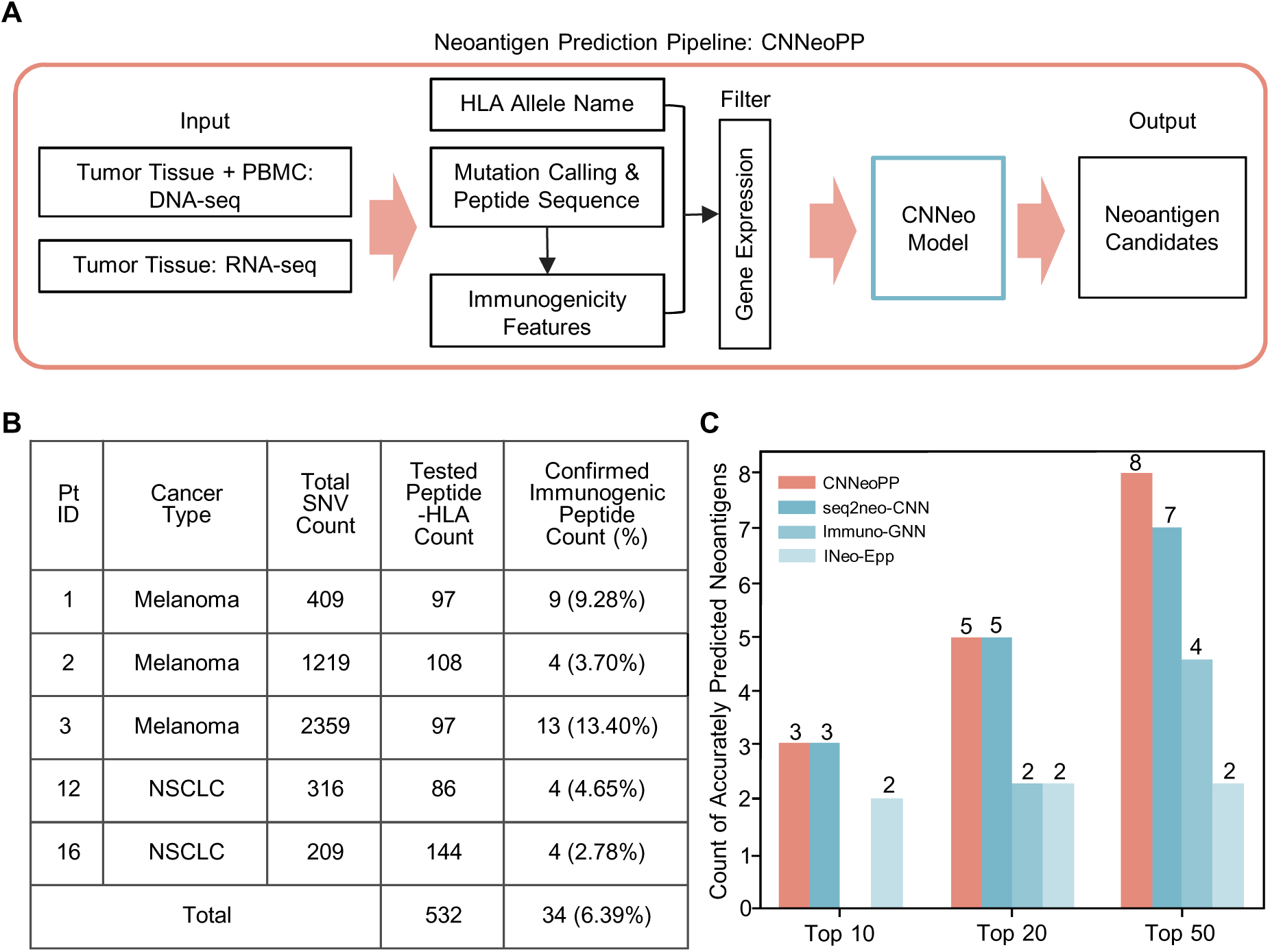
Independent Validation and Performance Assessment of CNNeoPP. (A) Schematic workflow of CNNeoPP. DNA-seq and RNA-seq data are processed to identify somatic SNVs, HLA types, and gene expression levels. Mutations are translated into amino acid changes, and 11 immunogenicity features are computed. These inputs are used by CNNeo to generate neoantigen predictions. (B) Summary of five TESLA patients’data used for independent validation of CNNeoPP, including cancer type, total identified neoantigen candidates, number of experimentally tested peptides, number of confirmed immunogenic neoantigens, and the percentage of confirmed immunogenic neoantigens. (C) Comparative performance evaluation of CNNeoPP against pipelines incorporating existing tools (seq2neo-CNN, Immuno-GNN, INeo-Epp) using TopN ranking analysis in TESLA cohort.

### Experimental Validation by T-cell Assay

This study enrolled one breast cancer (BCa) and two lung cancer (LC1 and LC2) patients, from whom tumor tissue and PBMC samples were collected (Fig. 5A). CNNeoPP was applied to process sequencing data and predict candidate neoantigens. To optimize the selection of peptide candidates for experimental validation, 50 top-ranked neoantigens were chosen from CNNeoPP predictions, with reference to existing model predictions. Of these, approximately half (n=27) were exclusively identified by CNNeoPP, while the remaining half overlapped with predictions from existing models and were prioritized in the final selection process. ELISpot assays with HLA-matched healthy PBMCs (Fig. S5A) were conducted to evaluate whether these neoantigens could elicit a T-cell response (Figs. 5B & S5B). After removing outliers, the average spot count for each peptide was calculated by subtracting the negative control background (Fig. 5C). Among the 50 peptides selected for experimental validation, 29 (58%) demonstrated immunogenicity, including 24 (48%) with a weak T-cell response and 5 (10%) with a strong response (Fig. 5D, Table S8). Notably, CNNeoPP-only predicted peptides exhibited a higher positive rate compared to peptides co-predicted by CNNeoPP and other models, with 59.3% vs. 34.8% showing a weak response and 11.1% vs. 8.7% showing a strong response. These findings further highlight the enhanced performance of CNNeoPP, as demonstrated by the higher proportion of positive responses in ELISpot assays relative to other prediction approaches.

**Figure 5.**
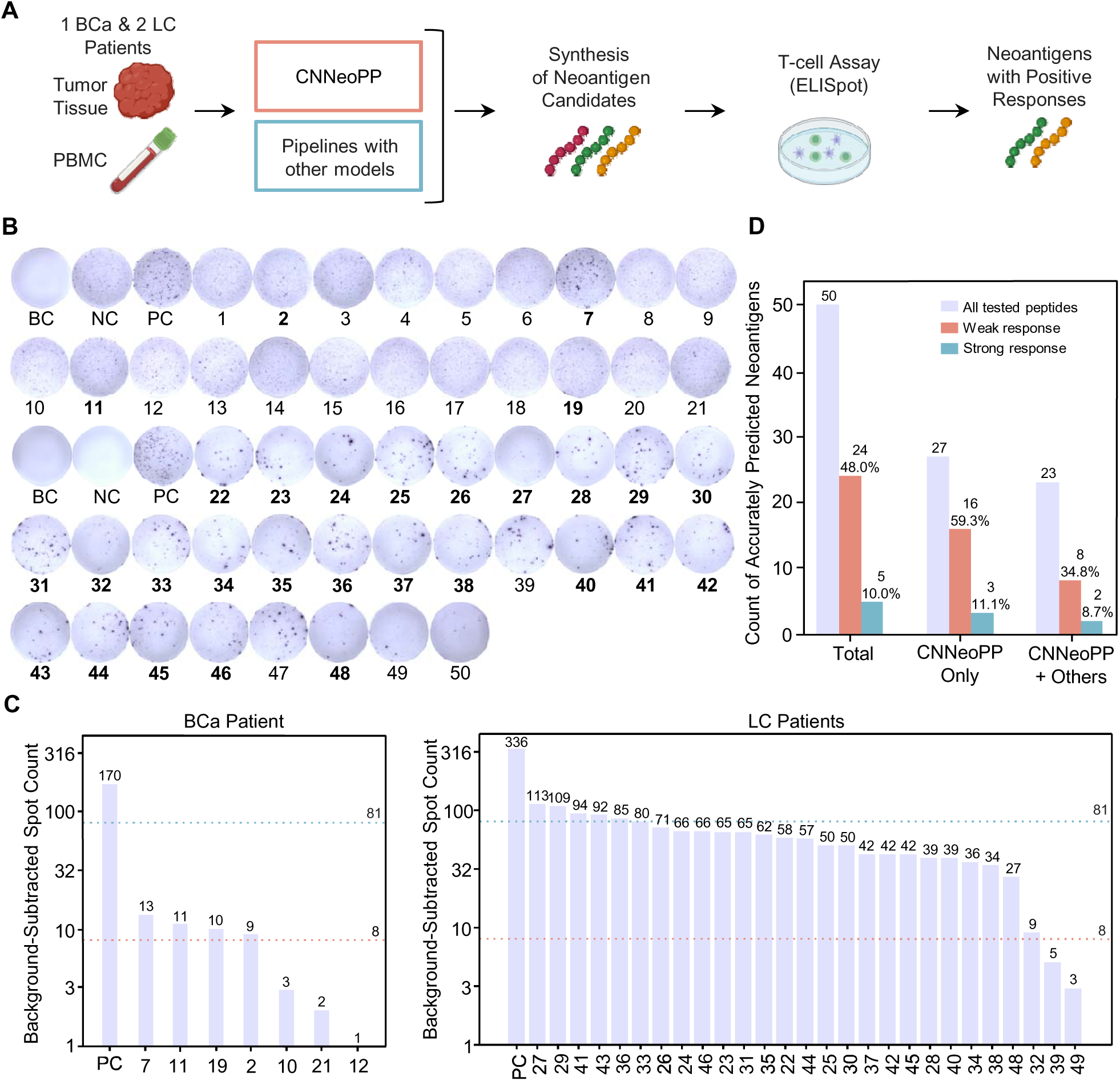
Experimental Validation of CNNeoPP via T-cell Assays. (A) Schematic workflow of experimental validation of CNNeoPP. Tumor tissue and PBMCs from breast and lung cancer patients were processed for DNA-seq and RNA-seq, followed by neoantigen prediction using CNNeoPP and other existing tools. A total of 50 selective neoantigen candidates were synthesized, and peptide-HLA complexes were tested for response in HLA-matched T cell assays. (B) Representative ELISpot results illustrate immune responses for blank control (BC), positive control (PC), negative control (NC), and tested neoantigen peptides. Peptides confirmed as immunogenic are highlighted in bold. (C) Background-subtracted IFN-_γ_ ELISpot responses for predicted neoantigens. Each bar represents a tested neoantigen, with PC exhibiting the highest response. The pink dashed line indicates the minimum response threshold (spot count _≥_ 8), while the teal dashed line represents the strong positive response threshold (spot count _≥_ 81). Neoantigens exceeding these thresholds were classified as immunogenic. (D) Performance evaluation of CNNeoPP, assessed by weakly positive (pink) and strongly positive (teal) responses across three groups: Total (50 peptides), CNNeoPP-only predicted peptides (27 peptides), and peptides predicted by combining CNNeoPP with other existing tools (23 peptides).

### Proof-of-Concept Study in cfDNA

To evaluate the feasibility of applying CNNeoPP to cfDNA-based neoantigen prediction, contrived empirical cfDNA was generated by mixing cancer cell line-derived sheared gDNA (sgDNA) with healthy plasma to achieve a controlled tumor content of 15% (Figs. 6A & S6A). Fragmentation analysis confirmed that the contrived cfDNA exhibited size distribution and fragmentation patterns consistent with natural cfDNA, ensuring its suitability as a representative cfDNA sample for feasibility validation of cfDNA-based prediction (Figs. 6B & S6B). After sequencing, CNNeoPP analyzed four different sample conditions: (I) a cancer cell line-derived sheared gDNA (sgDNA) sample with 100% tumor content at 200× coverage, (II & III) empirical cfDNA samples with 15% tumor content sequenced at 200× and 1000× coverage, and (IV) an *in silico* simulated cfDNA sample with 15% tumor content at 200× coverage (Fig. 6A).

**Figure 6.**
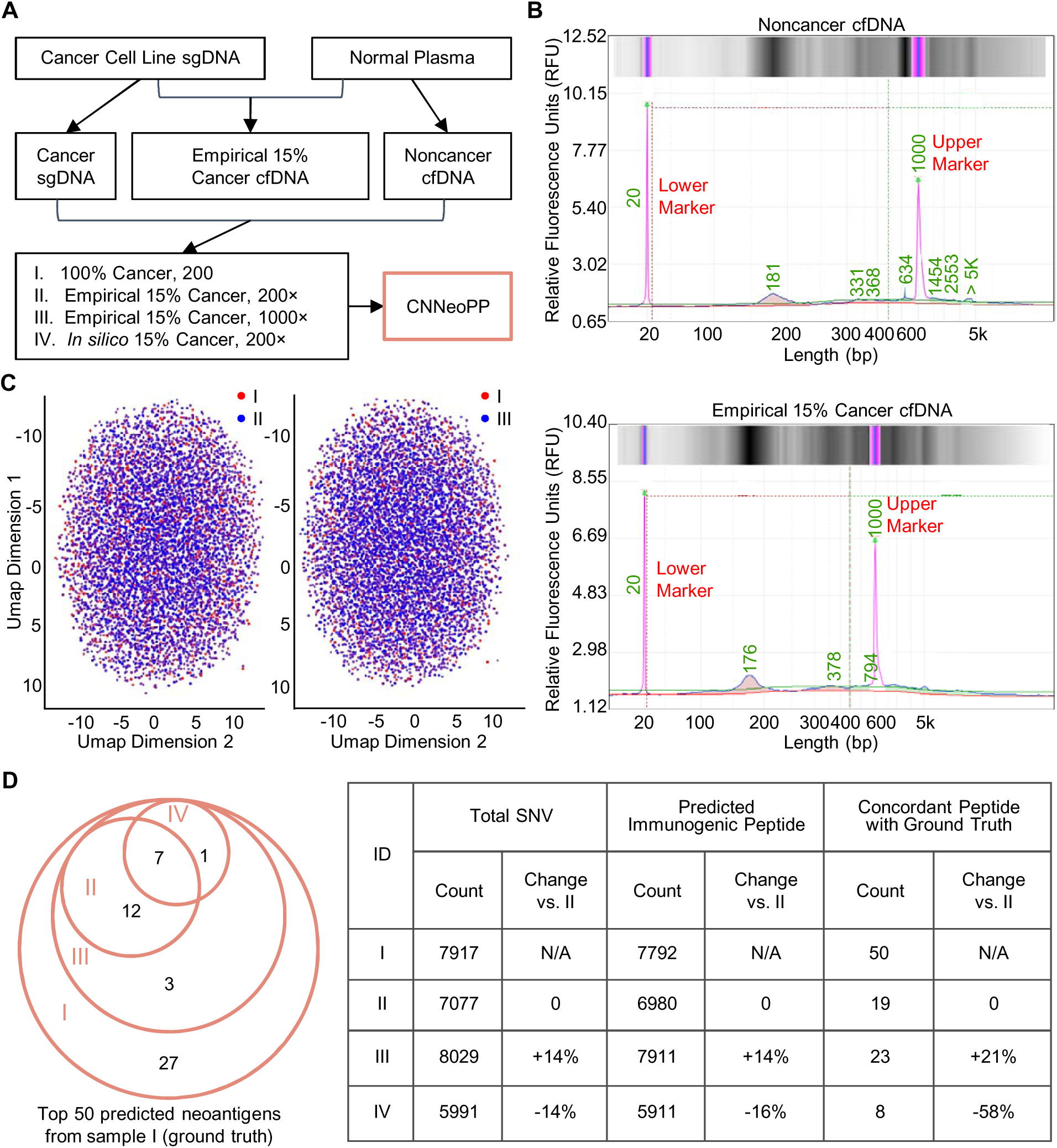
Proof-of-Concept Study for Neoantigen Prediction from cfDNA. (A) Experimental design for evaluating CNNeoPP performance from cfDNA. Cancer cell line-derived sheared gDNA (sgDNA) and non-cancer cfDNA were collected to generate an empirical contrived 15% cancer cfDNA sample, and all three were then sequenced at 200× coverage or additionally at 1000× coverage. Four conditions (I-IV) of samples were processed using CNNeoPP as annotated, including an *in silico* simulated 15% cancer cfDNA condition. (B) Fragmentation profiles of non-cancer cfDNA and empirical 15% cancer cfDNA samples. (C) UMAP visualization of 100% cancer sgDNA at 200× coverage and empirical 15% cancer cfDNA at 1000× and 200×, illustrating overlay patterns based on sequence features. Points are color-coded according to experimental conditions to highlight data distribution. (D) Summary of SNV and neoantigen predictions across different sample conditions. A Venn diagram (left) depicts the concordant prediction count across the other three conditions compared to the top 50 predicted neoantigens from condition I (100% cancer). The accompanying table (right) details total SNV counts, total predicted neoantigen counts, and the number of concordant neoantigens among the top 50 predicted from condition I, with the concordance percentage also reported.

UMAP visualization revealed similar clustering patterns between Sample I (tumor-only ground-truth scenario, red) and Sample II (contrived cfDNA, blue) (Fig. 6C, left). As the sequencing depth increased in Sample III, a higher concentration of blue dots appeared in the center of the distribution, leading to greater overlap (purple dots) and a reduction in non-overlapping red dots (Fig. 6C, right). Comparative analysis showed the *in silico* simulated 15% cancer sample (Sample IV) yielded a 16% (8 out of 50) overlap with “ground-truth” neoantigens as expected, whereas the empirical 15% cancer cfDNA sample (Sample II) demonstrated a higher 38% (19 out of 50) overlap and better clustering consistency with “ground-truth” (Figs. 6D & S7). Notably, increasing sequencing depth from 200× to 1000× (Sample II vs. III) led to a 14% increase in SNV calls and predicted neoantigens, while the “ground-truth” neoantigen prediction increased by 21% (Fig. 6D). These findings suggest that while neoantigen signals are weaker in cfDNA, this limitation may be partially compensated by increased sequencing depth, with CNNeoPP further amplifying this effect by identifying a higher proportion of “ground-truth” neoantigens beyond the expected increase in SNV detection.

### Establishment of CNNeoDB: A Neoantigen Database

To facilitate neoantigen research and predictive model development, a publicly accessible tumor neoantigen database CNNeoDB (http://www.cnneodb.cn/) was released. CNNeoDB integrates multi-source datasets that are used or reported in the present study, which allows users to search and download peptide entries based on criteria such as HLA allele, tumor type, and immunogenicity classification results. Researchers can contribute to CNNeoDB by submitting novel neoantigen candidates, including peptide sequences, genomic coordinates, and validation evidence.

## Discussion

A major challenge in advancing personalized cancer immunotherapy is the identification of individualized immunogenic neoepitopes, which remains a critical barrier to translating clinical studies into effective treatments. In this study, we developed and validated a comprehensive neoantigen prediction pipeline, CNNeoPP, that outperforms existing tools by accurately identifying neoantigens capable of eliciting T cell responses from individual patients. Furthermore, our proof-of-concept study revealed that while higher sequencing depth can partially recover suppressed neoantigen signals in cfDNA, additional strategies may be required for optimal detection.

Various physicochemical properties were found to differ significantly between immunogenic and non-immunogenic peptide sequences. Firstly, we demonstrated that 9-mers were the most abundant length in immunogenic peptides (59%), followed by 10-mers (28%), whereas the distribution was more comparable (49% vs. 46%) in non-immunogenic peptides. This result aligns with previous findings that 9-mer peptide binders are the most common binding peptides compared to 10-, 11-, and 8-mers across various cancer types and HLA alleles^14,15^. Additionally, our analysis highlighted unique residue preferences in immunogenic peptides, where negatively charged, hydrophilic residues (E, D) were enriched at positions P2, P3, P4, and positively charged residues (K, R) at position P1. In contrast, non-immunogenic peptides exhibited more generalized hydrophobic residues (e.g., L, V) distribution across all positions. While some aspects of our observations are novel, others align with previous studies indicating that neoantigen positions P2 and P3 tend to contain fewer hydrophobic residues^16^, and that negatively charged residues at P4 enhance binding affinity with TCR^17,18^. Furthermore, our data indicate that explicitly computed features are key contributors to immunogenicity classification, in agreement with previous findings^19,20^. Given the demonstrated importance of both the raw peptide sequence and explicitly computed features, both were incorporated as input data during CNNeo training and are expected to enhance its performance.

While various computational pipelines for neoantigen prediction have been developed, most focus predominantly on peptide-MHC binding affinity predictions. Recent approaches have started to incorporate additional features such as raw peptide sequences and HLA alleles to improve assessment of neoantigen immunogenicity^21,22^. Many existing neoantigen prediction tools and peptide analysis models still employ pre-deep-learning encoding methods, such as One-hot encoding (orthogonal encoding) ^23^. The key innovation of CNNeo lies in its specialized architecture, which uniquely integrates peptide sequence data using advanced NLP embedding techniques with structured immunogenicity features, enabling a more comprehensive and accurate neoantigen prediction framework. BioBERT, a pre-trained protein language model, can be fine-tuned for biological sequences^24^, while TF-IDF, a traditional NLP embedding method, quantifies the importance of sequence elements^25^. Though rarely applied to peptide analysis for neoantigen prediction, their integration with modern machine learning models offers a novel approach^26^. Convolutional layers in modern deep learning models have been extensively used to predict protein function from amino acid sequences^27^. FCNNs utilize dense layers to learn high-dimensional representations and improve classification tasks by consolidating both sequence-derived and additional structured data^28^. CNNeoPP leverages NLP-driven sequence processing, CNN-based feature extraction, and FCNN-based classification to synthesize both unstructured sequence data and structured features, maximizing neoantigen prediction accuracy.

Our study demonstrates the robustness of CNNeo and CNNeoPP through development with a diverse training set and comprehensive validations across independent datasets and experimental assays. CNNeoPP was developed using multiple curated datasets, ensuring broad applicability across different cancer types and patient HLA backgrounds^29,30^. CNNeoPP consistently outperforms existing tools^29,31–33^, maintaining high predictive accuracy across a series of ranking thresholds (top 10, 20, and 50), highlighting its reliability in prioritizing immunogenic candidates. Both model- and pipeline-level validation confirm its superiority, with CNNeo’s deep learning framework outperforming state-of-the-art prediction tools. In the TESLA validation dataset^1^, CNNeoPP underscored its performance by identifying 23.53% (8 out of 34) of the immunogenic peptides, while only requiring a total candidate selection rate of 9.40% (50 out of 532) within the dataset.

Beyond computational validation, IFN-γ ELISpot assays confirmed that neoantigens uniquely predicted by CNNeoPP exhibited a higher proportion of strong T cell responses compared to those identified by other tools. In the TESLA dataset, the true positive rate was 16% (8/50) across 5 patients. In contrast, in the experimental validation (3 patients), the positive rate was 58% (29/50) using predictions from CNNeoPP and other tools, based on a standard ELISpot threshold of 8 spots per well^34^. This rate further increased to 70% (19/27) for peptides exclusively predicted by CNNeoPP. When applying a more stringent threshold of 81 spot-forming cells per 4 × 10^4^ PBMCs^35^, the positive rate remained at 11.1% for CNNeoPP predictions, exceeding the 8.7% observed for peptides predicted by CNNeoPP and other tools combined. Notably, this 11.1% positive rate closely aligns with findings from a recent clinical trial on individualized mRNA vaccines encoding neoantigens that 11% of tested neoantigens induced a high-magnitude T cell response^36^. During the design and development of CNNeo, immunogenicity features relevant to TCR recognition, such as “Hydrophobicity of TCR contact residues”, were systematically integrated to enhance prediction accuracy, contributing to the high experimental validation positive rate observed here^1,37–39^. These findings underscore CNNeoPP’s ability to integrate features reflective of T cell recognition, establishing it as a powerful framework for guiding clinically relevant neoantigen selection.

Identifying neoantigens from plasma cfDNA offers a non-invasive, real-time approach to track tumor evolution and guide personalized immunotherapy and immune monitoring. High-depth sequencing is essential to improve sensitivity due to the significantly lower tumor content in cfDNA. Typically, only a small subset of somatic mutations (around 1-2%) give rise to neoantigens capable of eliciting T-cell responses^40–43^. Therefore, our proof-of-concept study evaluated the feasibility of CNNeoPP in detecting highly-ranked SNVs (top 0.64%, 50/7792) from a cell line-contrived sgDNA sample. Our results demonstrate that higher sequencing depth can partially recover suppressed neoantigen signals in cfDNA as identified by CNNeoPP. Moreover, our findings indicate that when analyzing high-depth samples, CNNeoPP further enhances this effect by prioritizing top-ranked neoantigens beyond the expected increase in SNV detection. Together, these results provide proof-of-concept evidence supporting the use of CNNeoPP for cfDNA-based neoantigen prediction. However, further optimization of library preparation is needed to maximize sensitivity and ensure robust detection of circulating neoantigens in liquid biopsy applications.

## Conclusion

This study presents CNNeo and CNNeoPP, a deep learning-based model and computational pipeline that significantly enhance neoantigen prediction accuracy. Through extensive validation using independent datasets and T-cell assays, CNNeoPP outperformed existing tools in identifying immunogenic neoantigens, with experimental results confirming higher T-cell response rates. Furthermore, our proof-of-concept study established the feasibility of cfDNA-based neoantigen prediction, supporting its potential for non-invasive tumor monitoring and personalized immunotherapy applications.

## Methods

### Study Participants and Sample Preparation

The study was approved by the Medical Ethics Review Committee of Northwest University with the approval number 24093085 and conducted per the Declaration of Helsinki. Breast cancer samples, including tumor tissue and PBMCs, were collected from four patients at Xi’an NO.3 Hospital, the Affiliated Hospital of Northwest University, while lung cancer samples from two patients were obtained from our biobank, as described in our previous study^44^. HLA-matched PBMCs were sourced from Milestone Biological Science & Technology Co., Ltd (Shanghai, China). Plasma samples were collected from healthy donors recruited at Northwest University. Genomic DNA (gDNA) and RNA from tumor tissues (either frozen or paraffin-embedded) and PBMCs were extracted using commercially available isolation kits (Tiangen Biotech Co., Ltd., Beijing, China). Noncancer cell-free DNA (cfDNA) was extracted from healthy plasma using the Magnetic Serum/Plasma DNA Kit (Tiangen Biotech Co., Ltd.) and quantified using a Qubit 4.0 Fluorometer (Thermo Fisher, Waltham, MA, USA). The fragment size of cfDNA was assessed using the Qsep 100 system (Bioptic Inc., Taiwan, China).

### Construction of Training and Validation Datasets

Paired HLA-peptide data used for training were curated and consolidated from published literature and existing neoantigen databases to develop predictive models (Fig. S1). A comprehensive search through Web of Science and PubMed identified studies reporting immunogenic and non-immunogenic tumor-specific peptides. Data extraction included mutated peptide sequences, wild-type sequences, HLA types, SNV mutation details, gene names, peptide lengths, and immunogenicity validation methods. Only peptides of 8-11 amino acids were included, while incomplete or duplicate entries were excluded. Additionally, neoantigen data were retrieved from Neodb and NEPdb databases (http://nep.whu.edu.cn). While Neodb contains only immunogenic peptides, NEPdb includes both immunogenic and non-immunogenic peptides. Each entry was cross-verified with the original publications to ensure accuracy. An independent dataset from a published study^45^, containing experimentally validated neoantigens and HLA pairs from 12 advanced lung cancer patients, was used for CNNeo model validation. Additionally, the TESLA dataset^1^, which includes raw DNA-seq and RNA-seq data from melanoma and NSCLC patients, along with experimentally validated neoantigens, was utilized for CNNeoPP pipeline validation. All four breast cancer and two lung cancer patients collected in-house were processed using CNNeoPP, with one breast cancer and two lung cancer samples selected for experimental validation. All training datasets, validation datasets, and in-house collected samples and data were consolidated into the CNNeoDB database.

### Cancer Cell Line and Contrived Empirical cfDNA Sample

The A549 lung cancer cell line was obtained from Procell Biotechnology Co., Ltd. (Wuhan, China) and cultured in RPMI-1640 medium supplemented with 10% FBS at 37°C in a 5% CO_2_ incubator. gDNA was extracted from A549 cells using the TIANamp Genomic DNA Kit (Tiangen Biotech Co., Ltd.). DNA fragmentation was performed using NEBNext® dsDNA Fragmentase® (New England Biolabs, Ipswich, MA, USA) to generate fragments of approximately 170 bp. The sheared gDNA (sgDNA) was then purified (size selection) using AMPure XP beads (Beckman Coulter Inc., Brea, CA, USA). To optimize yield, the incubation time was optimized to 40 minutes, and the AMPure XP bead-to-sample ratio was adjusted to 0.8× to selectively remove larger DNA fragments (Fig. S6A). To create an empirical 15% cancer cfDNA sample, the purified 100% cancer sgDNA was mixed with non-cancer cfDNA, resulting in a sample with approximately 15% tumor-derived DNA. The final DNA concentration was measured using the Qubit 4.0 Fluorometer (Thermo Fisher), and fragment size was assessed using the Qsep 100 system (Bioptic Inc.).

### Next Generation Sequencing

Whole-exome DNA sequencing (DNA-seq) and transcriptomic RNA sequencing (RNA-seq) of tumor tissue DNA, PBMC DNA, and tumor RNA were performed using the Illumina NovaSeq 6000 platform (Illumina, San Diego, CA, USA) at a sequencing depth of 100× coverage, with an input of 200/300 ng DNA/RNA per sample. DNA-seq of cell line-derived sgDNA and cfDNA samples, including the empirical 15% cancer cfDNA, was conducted on the MGI DNB-seq T7 platform (MGI Tech Co., Ltd., Shenzhen, China) at a sequencing depth of 200× or 1000× coverage, with an input of 50 ng DNA per sample.

### Feature Engineering and Evaluation

To identify key features for model training, a comprehensive literature review was performed. A total of 11 numerical features (F1-F11), termed as “immunogenicity features”, were identified as listed in Table S1. The calculations of these immunogenicity features are detailed in the Supplementary file. In addition to these structured data inputs, two unstructured data inputs including peptide sequences (F12) and HLA alleles (F13)^31^ were included. A key distinction in this study is the incorporation of HLA gene names (such as HLA-A*01:01), in addition to HLA pseudo sequences, for enhanced representation. To evaluate the independence of immunogenicity features, Spearman correlation analysis was conducted between each feature pair within the consolidated training dataset (n=1498). Relative feature importance was determined using a random forest classifier, where importance scores were computed to rank the feature accordingly.

### Structured Normalization and Sequence Embedding

To ensure uniform data distribution, Z-score normalization was applied to structured features using scikit-learn’s StandardScaler. This transformation centers the data at a mean of 0 and scales it to a standard deviation of 1. The scaler was fitted on the training dataset and applied consistently across both training and testing datasets to maintain data integrity throughout the analysis. To convert HLA names (or pseudosequences) and peptide sequences into numerical representations, NLP techniques were employed including one-hot encoding for categorical binary representation, TF-IDF (Term Frequency-Inverse Document Frequency) for quantifying sequence importance, and BioBERT, a pre-trained protein language model that tokenizes sequences and maps them to embeddings for downstream learning tasks. To mitigate class imbalance in the training dataset, SMOTE (Synthetic Minority Over-sampling Technique)^46^ was applied to generate synthetic samples for the minority class by interpolating between existing instances and their k-nearest neighbors.

### Development and Integration of Submodels into CNNeo and CNNeoPP

A total of 24 model combinations (Table S4) were trained using three encoders and four deep learning algorithms, optimized based on the characteristics of the input data and feature sets. Model training was conducted using Scikit-learn for traditional machine learning models and PyTorch for deep learning architectures. The dataset was split into training (60%), validation (20%), and test (20%) sets for deep learning models, while 5-fold cross-validation was applied for traditional machine learning algorithms. Typically, Random Forest (RF) and Fully Connected Neural Networks (FCNN) were utilized to process structured features (F1-F11), while Convolutional Neural Networks (CNN) were applied to encode unstructured features (F12, F13), which were transformed using NLP techniques, as described previously. The FCNN architecture consists of an initial fully connected layer, followed by a ReLU activation function, a dropout layer to prevent overfitting, and an additional fully connected layer, with classification performed using nn.CrossEntropyLoss, which applies softmax internally. The CNN architecture employs three parallel convolutional layers with kernel sizes of 3×3, 4×4, and 5×5, each followed by a ReLU activation function and max pooling to retain the most salient features. The outputs are concatenated, passed through a dropout layer to further reduce overfitting, and processed by a fully connected layer for classification. This parallel convolutional design enables the model to effectively capture multi-scale patterns within the data.

Three top-performing submodels were integrated into CNNeo model (Fig. 3B): 1) FCNN_TF: Peptide sequences and HLA alleles were encoded using TF-IDF and input into an FCNN. 2) CNN_BioBERT: A pre-trained BioBERT embedding was fine-tuned to encode paired mutation peptide sequences and HLA alleles, which were then processed through a CNN architecture. 3) FCNN_BioBERT: Both structured and unstructured features were integrated using multimodal fusion, where BioBERT tokenization was applied to unstructured data before merging it with structured features for training within an FCNN. Subsequently, CNNeo was integrated into CNNeoPP, a comprehensive neoantigen prediction pipeline, detailed in the Supplementary File. The performance of CNNeo and CNNeoPP was evaluated using independent datasets, as described above. The assessment was benchmarked against four existing neoantigen prediction tools: Seq2Neo-CNN^31^, Immuno-GNN^29^, DeepImmuno-CNN^32^, and INeo-Epp ^33^. Each tool was tested using default parameters, and a unified evaluation metric (Top N) was applied to ensure fair and consistent comparison.

### Experimental Validation by IFN-γ T-cell Assays

The clinical utility of CNNeoPP was evaluated using breast and lung cancer samples for neoantigen prediction. The IFN-γ ELISpot assays were performed by Baizhen Biotechnologies (Wuhan, China), an external research service provider specializing in immunological assays. The GenScript Biotech Corporation (Nanjing, China) was responsible for peptide synthesis and Baizhen Biotechnologies was responsible for assay setup, execution, and data acquisition following standardized protocols. The experimental workflow included PBMC QC and preparation, peptide synthesis and peptide stimulation, ELISpot plate processing, and spot count analysis, as outlined below. The top-ranked HLA-neoantigen pairs were compared with existing models, selecting 27 CNNeo-exclusive candidates and 23 overlapping candidates, totaling 50 neoantigens for validation. Peptides (95% purity) were synthesized by Milestone Biological Science & Technology Co., Ltd. and tested using HLA matched PBMCs in an IFN-γ ELISpot (Enzyme-linked immunospot) assay. Frozen PBMCs were rapidly thawed, washed to remove DMSO, and resuspended in complete medium. After cell counting, PBMCs were rested for 1 hour in a 37°C, 5% CO_2_ incubator, then 4×10^4^ cells per well were seeded pre-blocked ELISpot plates. Peptides were added at a final concentration of 20 µg/mL, alongside blank (RPMI-1640), positive (CEF peptide pool), and negative (RPMI-1640 with 0.5% DMSO and 10% FBS) controls. CEF is a control peptide pool derived from Cytomegalovirus (CMV), Epstein-Barr virus (EBV), and Influenza virus. Plates were incubated for 16–20 hours, washed, and incubated with biotin-labeled detection IFN-γ antibodies for 2 hours at room temperature, followed by streptavidin-horseradish peroxidase (HRP) incubation for 1 hour. 3,3’,5,5’-tetramethylbenzidine (TMB) substrate (Sigma-Aldrich, PA, USA) was applied for color development, and the reaction was stopped using deionized water. Plates were dried overnight in the dark before spot counts were recorded using an ELISpot reader. A weak positive response was defined as 8-81 spots after subtracting the background mean spot count, while a strong positive response was classified as ≥ 81 spots^34^. This threshold was derived from the 75th percentile of the spot count distribution, ensuring that strong responses were distinguished from weaker, non-significant responses in the dataset.

### cfDNA Sample Preparation and Sequencing Strategy

To evaluate the feasibility of CNNeoPP for cfDNA-based neoantigen prediction, four different sample types were prepared (Fig. 6A). A tumor-only sample (100% cancer sgDNA) was generated from cancer cell-derived sgDNA and sequenced at 200× depth. Two empirical 15% cancer cfDNA samples, containing 15% tumor content, were prepared by spiking 100% cancer sgDNA with noncancer cfDNA at a 15:85 ratio, followed by sequencing at 200× and 1000× depth, respectively. Lastly, an *in silico* simulated 15% cancer cfDNA sample was created by computationally mixing 100% cancer sgDNA reads (200× coverage) with noncancer cfDNA reads at the same 15:85 ratio, generating a simulated tumor cfDNA dataset. CNNeoPP was then applied to all samples to predict candidate neoantigens.

### Construction of CNNeoDB

To establish CNNeoDB, neoantigen data were compiled from multiple sources and categorized into two main datasets. Dataset 1 integrates (1) a publicly available dataset including training data for model development (Fig. S1A), (2) two validation datasets including the lung cancer dataset for CNNeo validation (Fig. S1B) and the TESLA dataset for CNNeoPP validation (Fig. S1C), and (3) one new experimentally validated neoantigens dataset developed from three cancer patients in this study. Dataset 2 consists of candidate neoantigens identified by CNNeoPP in additional cancer patients. These datasets provide a comprehensive resource for neoantigen prediction and validation.

### Statistical Analysis

All statistical analyses and data visualization were performed using Python-based libraries. Spearman correlation analysis was applied to assess relationships between immunogenicity features using correlation coefficients, while chi-square tests evaluated differences in peptide length distributions between immunogenic and non-immunogenic peptides. Shapiro-Wilk tests were conducted to assess whether immunogenic features followed a normal distribution, revealing that most features did not meet normality assumptions (Table S3). Consequently, Mann-Whitney U tests were applied to compare the distribution of immunogenicity features across peptide groups. P-values were reported where applicable to indicate statistical significance.

## Data Availability

All data produced in the present study are available in the manuscript, supplementary materials, or through the CNNeoDB database (http://www.cnneodb.cn/). Additional data may be available upon reasonable request to the authors.

## Acknowledgements

This work was supported by the National Natural Science Foundation of China (81672627, 82071863). This research was supported by Global - Learning& Academic research institution for Master’s/PhD students, and Postdocs (LAMP) Program of the National Research Foundation of Korea (NRF) grant funded by the Ministry of Education (No. RS-2024-00443714). We sincerely acknowledge and express our gratitude to the patients and donors for their invaluable contributions of samples and data. Their generosity has been instrumental in advancing research on neoantigen prediction and personalized cancer immunotherapy.

## Author contributions

Y.C. and X.Z. developed and directed the total project. Y.C. executed all the analyses, including data preprocessing, model development, model implementation, experiment and downstream analyses. R.C. contributed to the partial data preprocessing and model development. Y.C., R.C., L.W. and X.Z. contributed to the interpretation of the model’s results. Y.G.L., Y.L.C. and X.Z. contributed to the experimental design. M.M.S., Z.R.H., D.Y.Y., S.T.Z. and S.H.G. contributed to the experiment. Y.C. and X.Z. generated the figures and tables, as well as wrote, edited and revised the manuscript with input from all authors. S.H. and L.B. contributed to manuscript review and statistical analysis oversight. X.Z. contributed to study design, data analysis oversight and funding acquisition.

---

**Supplementary Figure S1.**
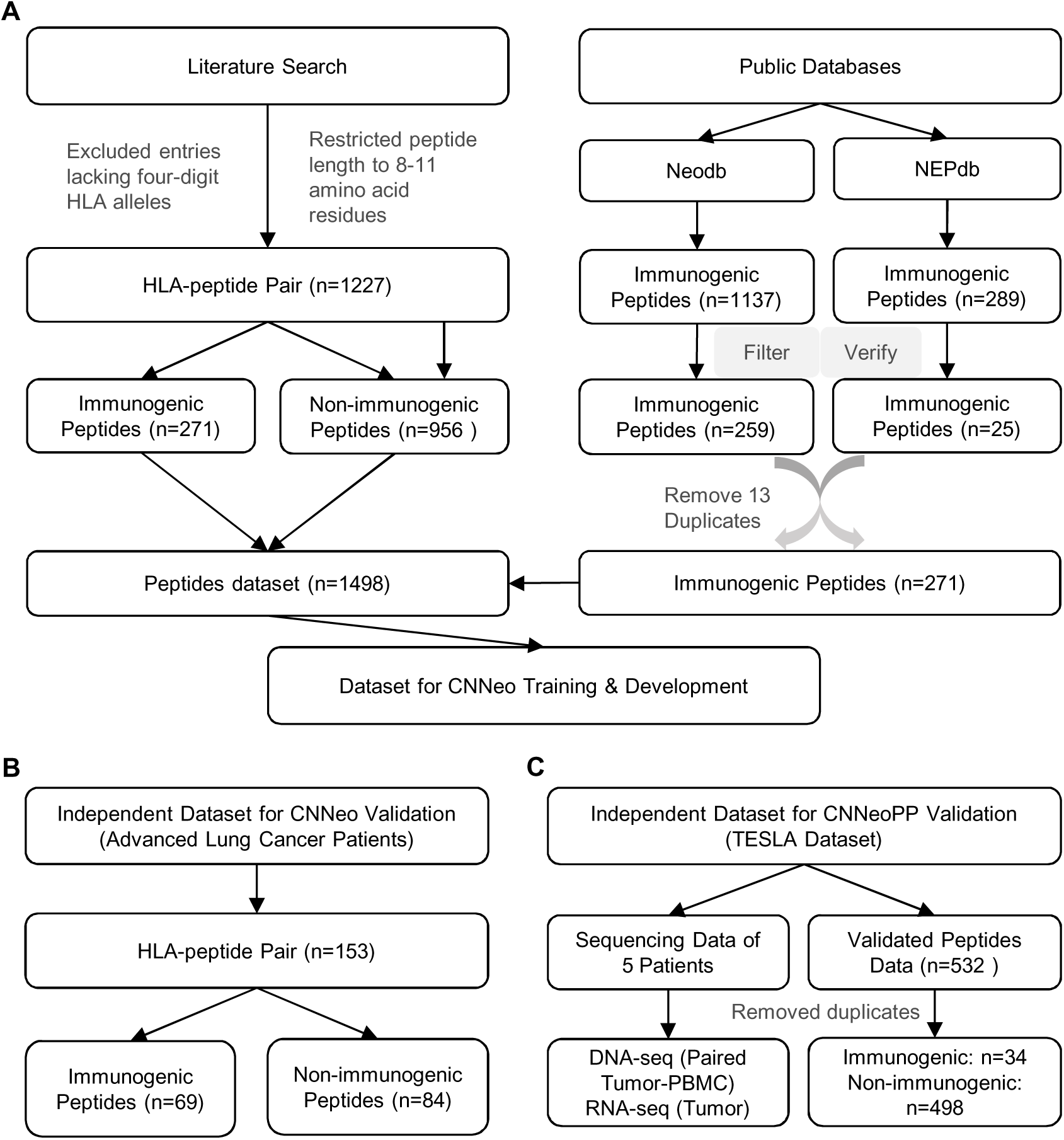
Workflow of public dataset collection and consolidation in this study. (A) Datasets used for CNNeo training and development (Training dataset). (B) Independent dataset for CNNeo validation (Advanced lung cancer patients). (C) Independent dataset for CNNeoPP validation (TESLA dataset).

**Supplementary Figure S2.**
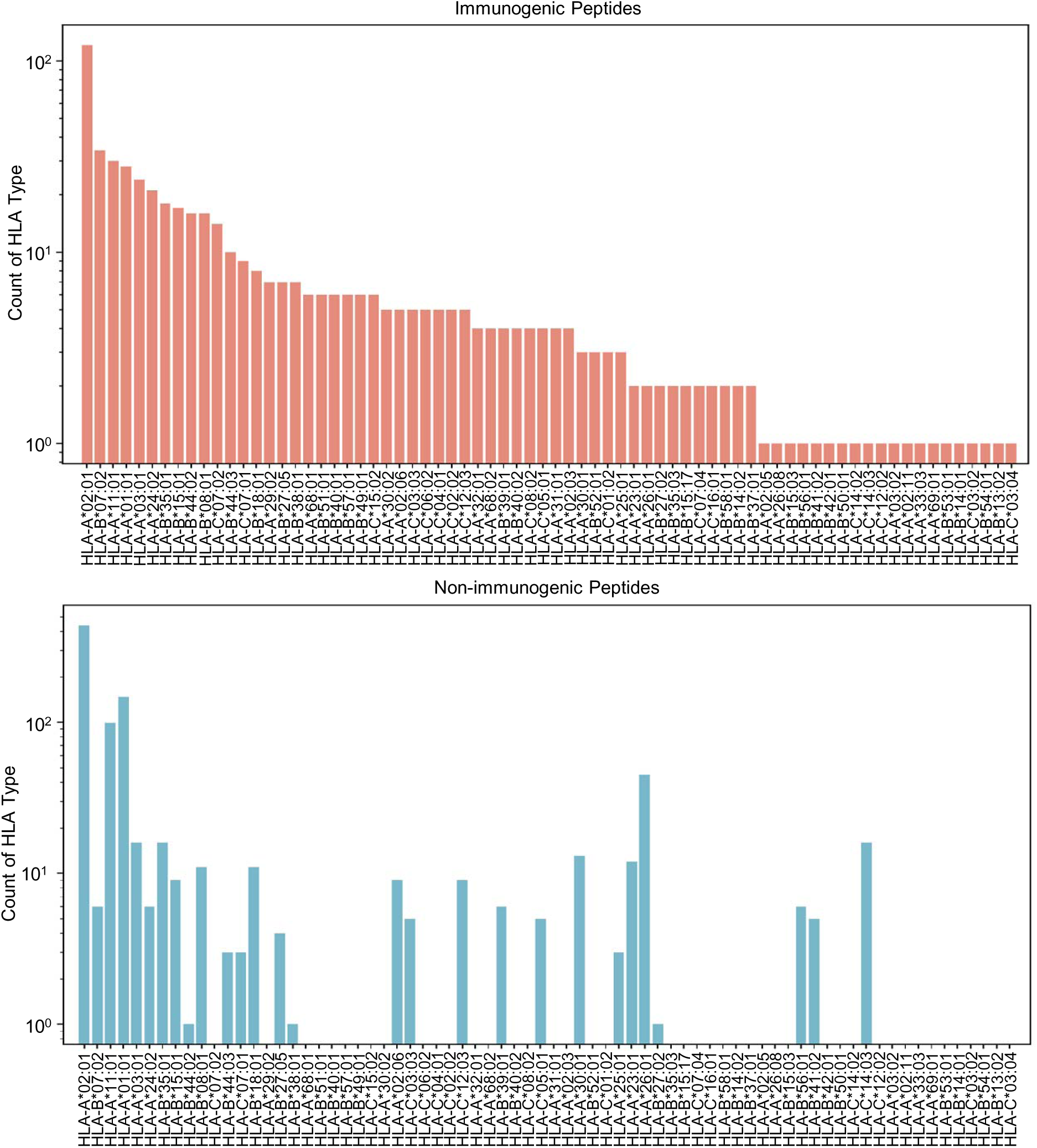
Distribution of HLA genotypes paired with immunogenic (upper) and non-immunogenic (lower) peptides from Training dataset.

**Supplementary Figure S3.**
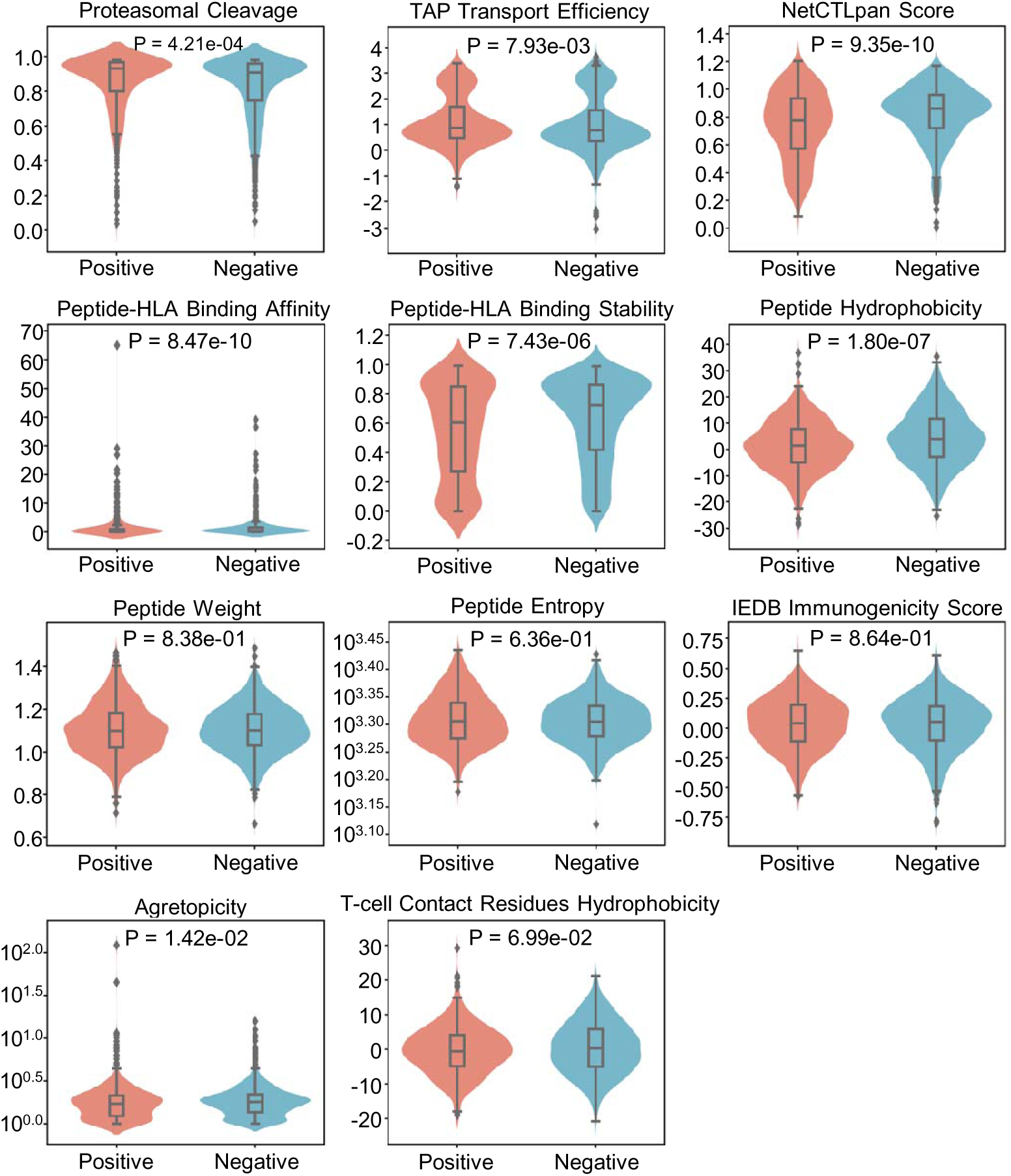
Comparative visualization of 11 immunogenicity features between immunogenic (Positive) and non-immunogenic (Negative) datasets. Violin plots represent kernel density estimation, where wider sections indicate higher probability density. Box plots within the violins display the second and third quartiles with the interquartile range (IQR), with the median shown as a line inside the box. Whiskers extend to the first and fourth quartiles ± 1.5 × IQR or the minimum/maximum values within this range. Statistical significance was assessed using the Mann-Whitney U test, with p-values indicated in the plots.

**Supplementary Figure S4.**
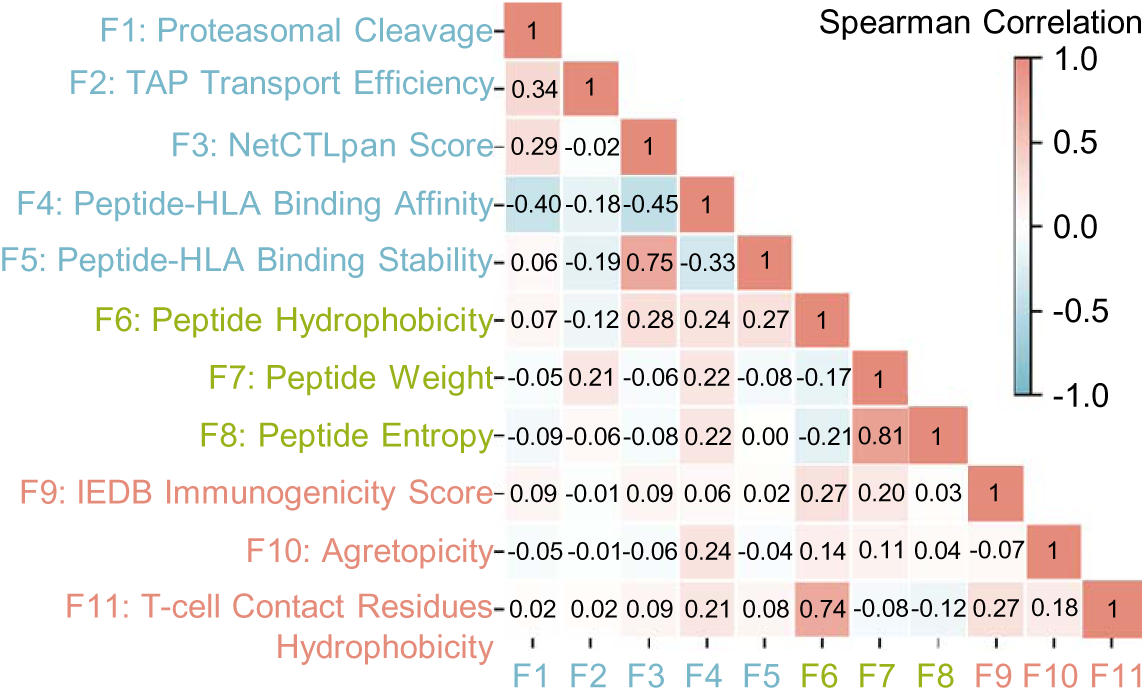
Spearman Correlation Coefficient matrix of 11 immunogenicity features used in CNNeo. The color intensity reflects the absolute value of the Spearman Correlation Coefficient, indicating the strength of positive or negative correlations. Feature names are color-coded to indicate their respective categories: Neoantigen processing and presentation (F1-F5), Biochemical properties of neoantigens (F6-F8), and T cell recognition (F9-F11).

**Supplementary Figure S5.**
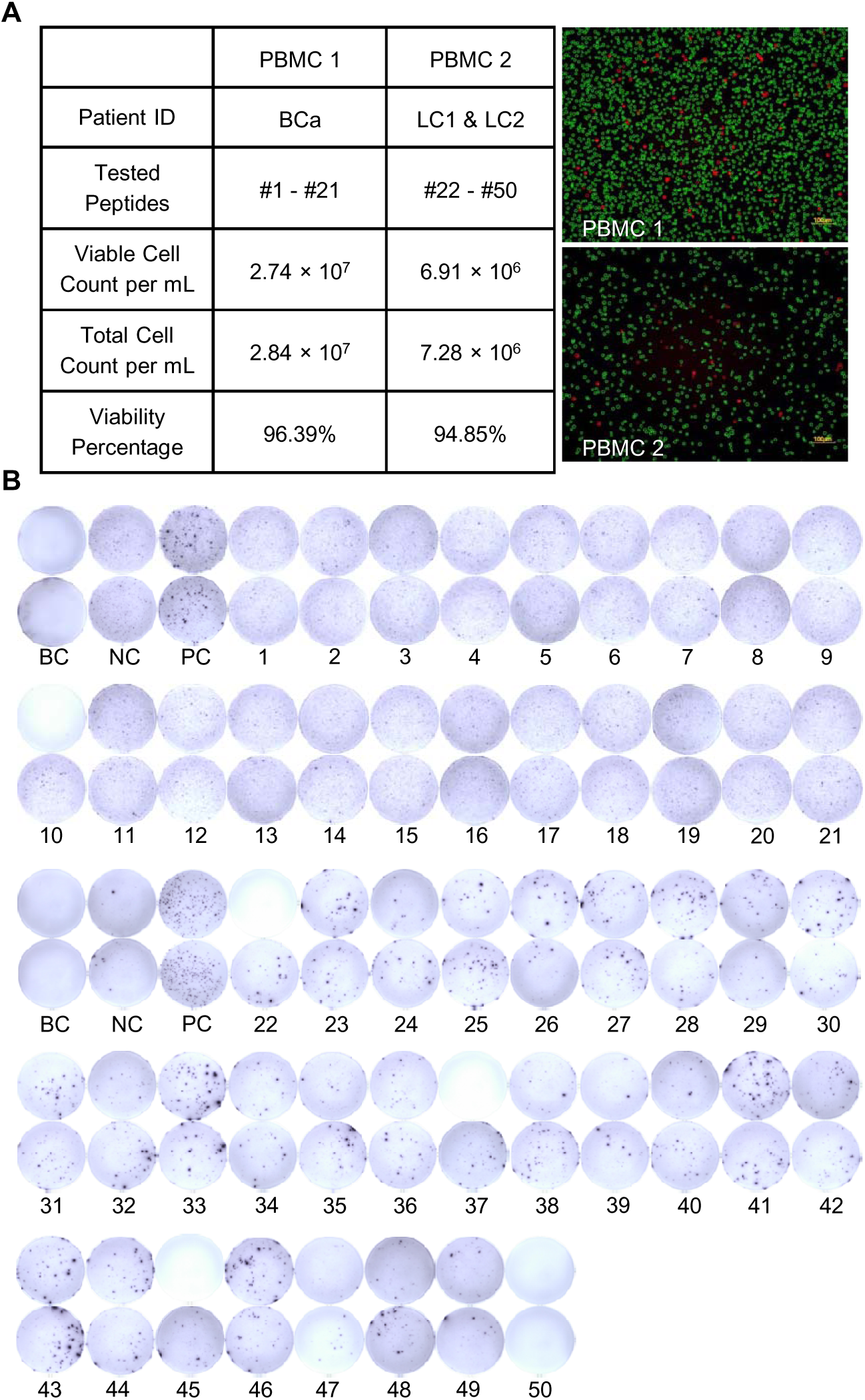
Quality Control of PBMCs and ELISpot Validation of Neoantigen Responses. (A) Quality control assessment of PBMCs used in the experimental validation of CNNeoPP. A QC table is included, summarizing the total cell count, viable cell count, and cell viability percentage. Live/dead staining of PBMCs is shown, where green represents live cells and red represents dead cells. (B) ELISpot validation of neoantigen responses in three cancer patients. Additional replicate images are provided to complement the representative results shown in Figure 5B. Blank control (BC), positive control (PC) and negative control (NC) groups were included.

**Supplementary Figure S6.**
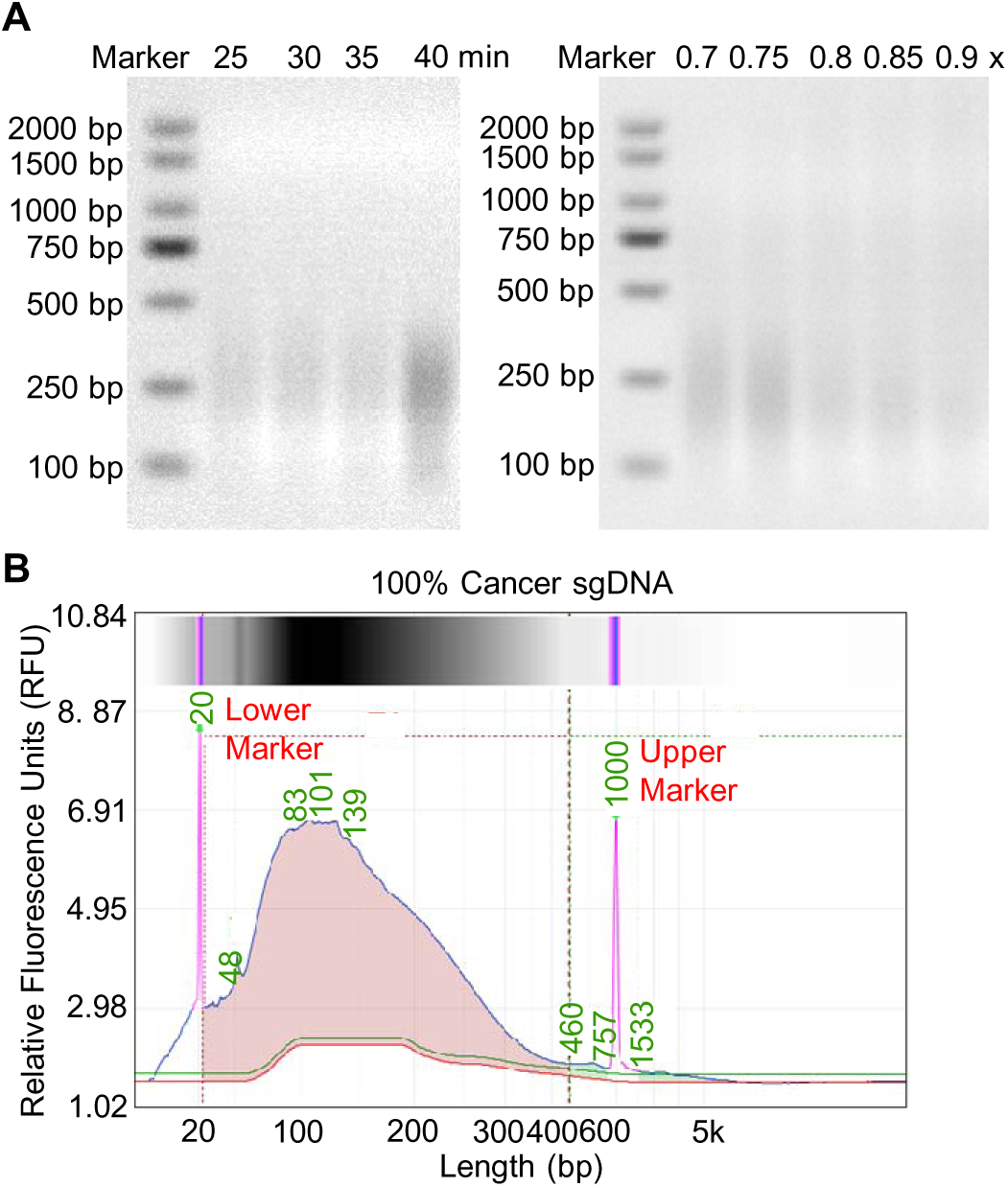
Optimization tests for the preparation of cell-line derived cancer sgDNA samples. (A) Agarose gel electrophoresis comparing DNA fragment sizes across various enzymatic digestion time points (left) and during size selection using different AMPure XP bead-to-sample volume ratios as annotated (right). The optimized conditions were determined to be 40 minutes of digestion and a 0.80× volume ratio. (B) Fragmentation profiles of sheared genomic DNA (sgDNA) derived from cancer cell line.

**Supplementary Figure S7.**
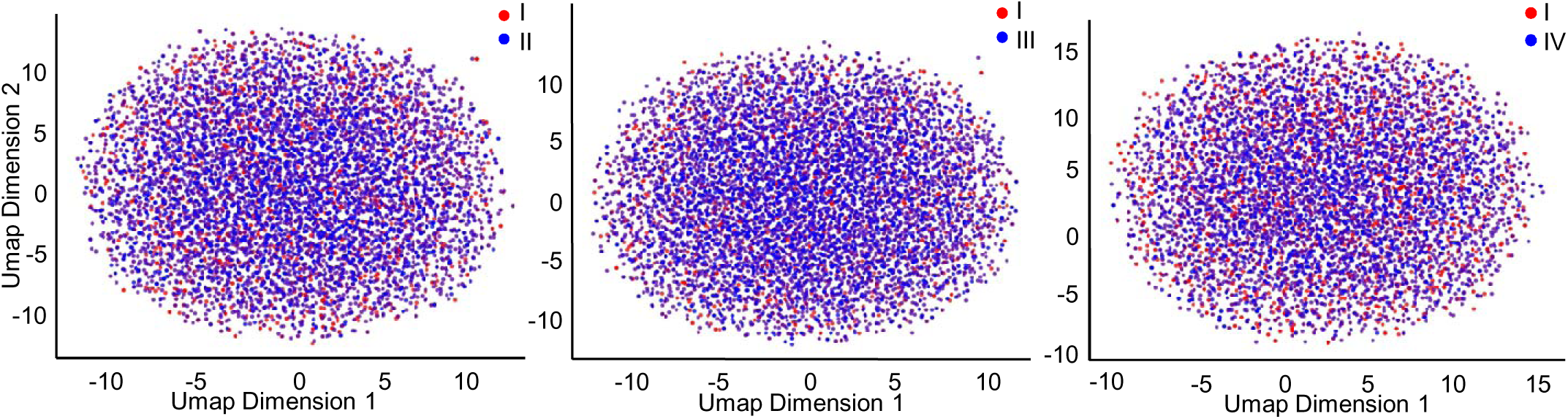
UMAP visualization and comparison between 100% cancer sgDNA at 200× coverage with empirical 15% cancer cfDNA at 1000× and 200×, as well as in silico 15% cancer cfDNA at 200×, illustrating overlay patterns based on sequence features. Points are color-coded according to experimental conditions to highlight data distribution.

**Table S1.** Summary of literature-sourced datasets used for CNNeo training and development.

**Table S2.** List of input features used for CNNeo training and development.

**Table S3.** Normality test for the distribution of 11 immunogenicity features in the Training dataset.

**Table S4.** Performance metrics of all individual models evaluated for CNNeo development.

**Table S5.** List of peptides predicted by CNNeo and existing tools in an independent dataset.

**Table S6.** List of raw sequencing files in TESLA dataset.

**Table S7.** List of peptides predicted by CNNeoPP and existing tools in TESLA dataset.

**Table S8.** List of peptides tested and predicted in experimental validation of CNNeoPP.

