## Supplementary File for "CNNeoPP: A Deep Learning Pipeline for Personalized Neoantigen Prediction and Liquid Biopsy Applications"

**Derivation of Immunogenicity-Associated Features**

To define features associated with neoantigen immunogenicity, a comprehensive literature review was conducted. Eleven numerical features (F1–F11) were identified and calculated for each peptide–HLA pair as described below:

Proteasomal Cleavage, TAP Transport Efficiency and NetCTLpan Score: These three features were calculated using NetCTLpan-1.1^1^, providing estimates of proteasomal cleavage likelihood, TAP transport efficiency, and a composite NetCTLpan processing score.

Peptide-HLA Binding Affinity: Binding affinities for mutant and wild-type peptides were predicted using NetMHCpan-4.1^2^, with results expressed as %rank values. Peptides with %rank < 0.5 were classified as strong binders, and those with %rank < 2 were defined as weak binders.

Peptide-HLA Binding Stability: Binding stability was predicted using NetMHCstabpan-1.0, and reported as predicted stability (Pred) values^3^.

Peptide Hydrophobicity: Hydrophobicity was calculated by summing the Kyte–Doolittle values of all amino acids within the mutant peptide sequence (AAindex JURD980101).

T-cell Contact Residues Hydrophobicity: Hydrophobicity scores were also calculated specifically for the T-cell contact region (residues at positions 4 to n–1, excluding anchor residues), using the same hydrophobicity scale.

Peptide Weight: The molecular weight of each peptide was determined by summing the monoisotopic masses of individual residues and subtracting the mass of water (18.015 Da) per peptide bond.

Peptide Entropy: Entropy scores were calculated by summing residue-specific values from AAindex HUTJ700103.

IEDB Immunogenicity Score: Immunogenicity scores were obtained using the IEDB Immunogenicity Prediction Tool, which evaluates amino acid composition at T-cell receptor contact positions (http:// tools.iedb.org/immunogenicity/).

Agretopicity: This metric was calculated as the ratio of binding affinity (IC50) between the mutant and corresponding wild-type peptide–HLA complexes, representing differential binding potential.

**Processing of DNA-seq and RNA-seq Data**

Whole-exome sequencing (WES) data underwent quality control using FastQC (v0.12.1)^4^. Adapter sequences and low-quality bases (Phred < 33) were removed using Trimmomatic (v0.39)^5^. Reads were aligned to the hg38 human genome using BWA (v0.7.12)^6^. Resulting BAM files were sorted using SAMtools, and PCR duplicates were removed with Picard’s MarkDuplicates. Base quality score recalibration (BQSR) was performed using GATK (v4.4)^7^. RNA-seq data were quality-filtered using Cutadapt, with adapters removed and bases with Phred scores < 25 trimmed to ensure high integrity for expression quantification.

**Somatic Mutation Calling and Filtering**

Somatic SNVs were identified using Mutect2 (GATK v4.4) in paired tumor–normal mode. BQSR-processed BAM files were used, and germline variants were filtered by applying a germline resource file. Cross-sample contamination was estimated using GetPileupSummaries and CalculateContamination, generating contamination tables for filtering. Mutations were filtered using FilterMutectCalls, and those labeled as “PASS” were retained with SelectVariants for downstream neoantigen prediction.

**Mutation Annotation and Peptide Extraction**

Somatic mutations were annotated using ANNOVAR, focusing on nonsynonymous variants. A custom Python (v3.9.12) script was used to translate each variant into a 21-mer mutant peptide sequence centered on the mutation, paired with the wild-type sequence. These were then fragmented into 8–11-mer peptides, ensuring the mutation was retained within each fragment to conform to MHC class I binding requirements.

**Gene Expression Quantification and Filtering**

Gene isoform expression levels were quantified using Kallisto^8^, with the reference transcriptome from Ensembl GRCh38. An index was generated for quantification, and expression was reported in transcripts per million (TPM). Only peptides derived from transcripts with TPM ≥ 1 were retained, ensuring biological relevance in downstream prediction

**HLA Genotyping and Binding Affinity Prediction**

HLA class I alleles were inferred from normal WES data using OptiType^9^, which achieves ~97% genotyping accuracy^10^. Binding affinities for mutant and wild-type peptides were predicted using NetMHCpan-4.1^2^. Peptides with a %rank > 2 were excluded to retain only strong and weak binders likely to be presented by HLA class I molecules.

**Immunogenicity Prediction Using CNNeo**

Candidate neoantigens, their HLA types, and all 11 computed features were input into the CNNeo model for immunogenicity prediction. Each peptide–HLA pair was assigned a numeric immunogenicity score, which quantitatively reflects the likelihood of eliciting a T-cell response. Higher scores indicated a greater predicted immunogenic potential. These scores were used to rank candidate peptides, enabling systematic prioritization for downstream analysis, validation, and integration into CNNeoPP.
